# Inpatient Hospitalizations for COVID-19 Among Patients with Prader-Willi Syndrome: a National Inpatient Sample Analysis

**DOI:** 10.1101/2024.09.06.24313191

**Authors:** James Luccarelli, Theresa V. Strong, Emily B. Rubin, Thomas H. McCoy

## Abstract

**Background:** Prader-Willi syndrome (PWS) is a genetic disorder associated with baseline respiratory impairment caused by multiple contributing etiologies. While this may be expected to increase the risk of severe COVID-19 infections in PWS patients, survey studies have suggested paradoxically low disease severity. To better characterize the course of COVID-19 infection in patients with PWS, this study analyzes the outcomes of hospitalizations for COVID-19 among patients with and without PWS.

**Methods:** The National Inpatient Sample, an all-payors administrative claims database of hospitalizations in the United States, was queried for patients with a coded diagnosis COVID-19 in 2020 and 2021. Hospitalizations for patients with PWS compared to those for patients without PWS using Augmented Inverse Propensity Weighting (AIPW).

**Results:** There were 295 (95% CI: 228 to 362) COVID-19 hospitalizations for individuals with PWS and 4,112,400 (95% CI: 4,051,497 to 4,173,303) for individuals without PWS. PWS patients had a median age of 33 years compared to 63 for those without PWS. Individuals with PWS had higher baseline rates of obesity (47.5% vs. 28.4%). AIPW models show that PWS diagnosis is associated with increased hospital length of stay by 7.43 days, hospital charges by $80,126, and the odds of mechanical ventilation and in-hospital death (odds ratios of 1.79 and 1.67, respectively).

**Conclusions:** PWS patients hospitalized with COVID-19 experienced longer hospital stays, higher charges, and increased risk of mechanical ventilation and death. PWS should be considered a risk factor for severe COVID-19, warranting continued protective measures and vaccination efforts. Further research is needed to validate coding for PWS and assess the impact of evolving COVID-19 variants and population immunity on this vulnerable population.

## Introduction

Prader-Willi syndrome (PWS) is a genetic disorder caused by loss of function of the paternally inherited 15q11-q13 locus [Chung et al., 2020]. Multiple genetic mechanisms are responsible for this imprinting disorder, which in total affects 1/10,000 to 1/30,000 births [Cassidy et al., 2012]. Clinical features of PWS are age-dependent, with wide-ranging neuromuscular, endocrine, and behavioral manifestations [Butler et al., 2018; Manzardo et al., 2018; Bohonowych et al., 2019]. Multiple of these coalesce to result in impaired pulmonary mechanics, respiratory control, and sleep-disordered breathing [Gillett and Perez, 2016].

Individuals with PWS are at high risk of developing obesity as a result of hyperphagia and food-seeking behaviors [Dykens et al., 2007], with subsequent mechanical effects on lung function [Peters and Dixon, 2018]. This is compounded by hypotonia, another core feature of PWS, which results in respiratory muscle weakness [Hákonarson et al., 1995]. Respiratory dysfunction in individuals with PWS occurs even in the absence of obesity, however, largely through central mechanisms. These include reduced chemoreceptor function in PWS [Gozal et al., 1994] resulting in aberrant ventilatory responses to hypercapnia, hypoxia, and hyperoxia independent from the degree of obesity [Schlüter et al., 1997; Livingston et al., 1995]. There is also substantial hypothalamic dysfunction in PWS [Tauber and Hoybye, 2021], with corresponding effects on respiration and sleep/wake function [Itani et al., 2023]. Individuals with PWS also have very high rates of obstructive sleep apnea and aspiration. Together these mechanisms are responsible for the high rates of respiratory disorders in individuals with PWS [Festen et al., 2006]. Respiratory disorders are also the greatest cause of death in individuals with PWS in longitudinal studies, accounting for 31-50% of deaths [Butler et al., 2017; Pacoricona Alfaro et al., 2019], with high rates of death due to both respiratory failure and respiratory infections.

In the general population, obesity is associated with greater severity of COVID-19 infection and worse outcomes [Russo et al., 2023; Gao et al., 2021; Kompaniyets, 2021], as is obstructive sleep apnea [Arish et al., 2023]. Moreover, COVID-19 infection itself may increase risk of aspiration [Grilli et al., 2022] and may directly cause respiratory muscle weakness [Severin et al., 2022], conditions which are highly prevalent in PWS patients at baseline. Given this, infection with the COVID-19 virus may be expected to be especially dangerous for individuals with PWS. Initial evidence, however, suggest paradoxically low severity of COVID-19 infections among such individuals. One survey in France between March 2020 and Jan 2021 of 288 adults with PWS found that 13.2% had confirmed COVID-19 infection, with 12 of the 38 infected individuals (31.6%) admitted to the hospital, with no ICU admissions and no deaths [Coupaye et al., 2021]. Among these hospitalized cases, however, 10 were residing in the hospital setting at time of infection. Among 239 children with PWS in the same study, 5.4% had COVID-19 during the study period with no hospitalizations or deaths. This infection rate was lower than that of the general French population at the time. A second international survey of 72 individuals with PWS from May 2020 through April 2021 identified 56 individuals diagnosed with COVID-19 (77.8%), of whom six (10.7%) were hospitalized, with no deaths [Whittington et al., 2022].

While these survey findings suggest that COVID-19 infections in individuals with PWS may be unexpectedly mild, such surveys are inherently limited by response bias and thus cannot inform disease severity on a population level. A greater understanding of the course of COVID-19 hospitalizations among individuals with PWS would help optimize clinical care of individuals with PWS and may point towards unique disease pathology in such patients which would provide information about COVID-19 infections in the broader population. This study analyzes COVID-19 infections among individuals with and without PWS in a large all-payor database of US hospital discharges in 2020 and 2021.

## Methods

### Data Source

This study analyzes the 2020 and 2021 editions of the National Inpatient Sample (NIS), an all-payors database of inpatient hospitalizations in the United States prepared by the Healthcare Cost and Utilization Project (HCUP) of the Agency for Healthcare Research and Quality. The NIS samples hospitalizations in non-federal acute care hospitals in 49 US states, with sampled hospitalizations weighted to produce national estimates and associated variances. This variance is presented to describe sampling uncertainty in the overall number of hospitalizations, whereas weighted point estimates are given for all other values. The NIS provides demographic details, information about hospital length of stay and overall hospital charges, and up to 40 discharge diagnoses for each hospitalization. This study was reviewed by the Mass General Brigham Institutional Review Board and classified as Exempt.

### Data Selection

Hospitalizations for individuals with PWS were those that included *International Statistical Classification of Diseases, Tenth Revision, Clinical Modification* (*ICD-10-CM*) code Q87.11 among the discharge diagnosis list [Luccarelli, 2022]. Encounters involving COVID-19 infection were defined as those with a discharge diagnosis of U07.1 (COVID-19). This code has been validated as having high sensitivity and positive predictive value for identifying COVID-19 cases [Wu et al., 2022; Kluberg et al., 2022]. Patients who received mechanical ventilation were identified based on ICD-10 procedural codes 5A1935Z, 5A1945Z, and 5A1955Z. Comorbidities were calculated using the AHRQ Elixhauser Comorbidity Software Refined for ICD-10-CM, as listed in the NIS Diagnosis and Procedure Groups File. Patients of all ages were included in the analysis.

### Statistical Analysis

Due to privacy restrictions for the NIS, subgroups containing fewer than 11 admissions are reported as “<11”, and no subgroup analyses are conducted which would require sample sizes <11. Demographics and diagnoses are presented using descriptive statistics using SPSS (version 29; IBM Software, Inc, Armonk, NY). For the primary statistical analysis, Augmented Inverse Propensity Weighting (AIPW) was used to estimate the effect of PWS diagnosis (yes/no) on the core outcomes of, receipt of mechanical ventilation (yes/no), in-hospital death (yes/no), hospital length of stay (days), and total hospital charges ($). AIPW utilizes doubly robust estimation, which combines outcome regression with weighting by the propensity score to provide an effect estimator that is robust to misspecification of either the outcome model or the propensity model [Funk et al., 2011]. PWS (yes/no) was the binary exposure variable in the models, with age, sex, race, primary hospital service line, and year as covariates. AIPW models utilized 10-fold cross validation. Calculations were performed using the AIPW package (v0.6.3.2)[Zhong et al., 2021] in R Statistical Software (v4.3.3; R Core Team 2021).

## Results

In 2020 and 2021 there were a total of 295 (95% CI: 228 to 362) hospitalizations for individuals with PWS involving infection with COVID-19 and 4,112,400 (95% CI: 4,051,497 to 4,173,303) hospitalizations involving a diagnosis of COVID-19 infection among individuals without PWS. Full demographic information is shown in Table 1. Patients with PWS hospitalized with COVID-19 had a median age of 33 years compared to a median age of 63 years for those without PWS (Figure 1). Males represented 62.7% of COVID-19 hospitalizations for individuals with PWS compared to 51.7% of COVID-19 hospitalizations for those without PWS.

**Figure 1:**
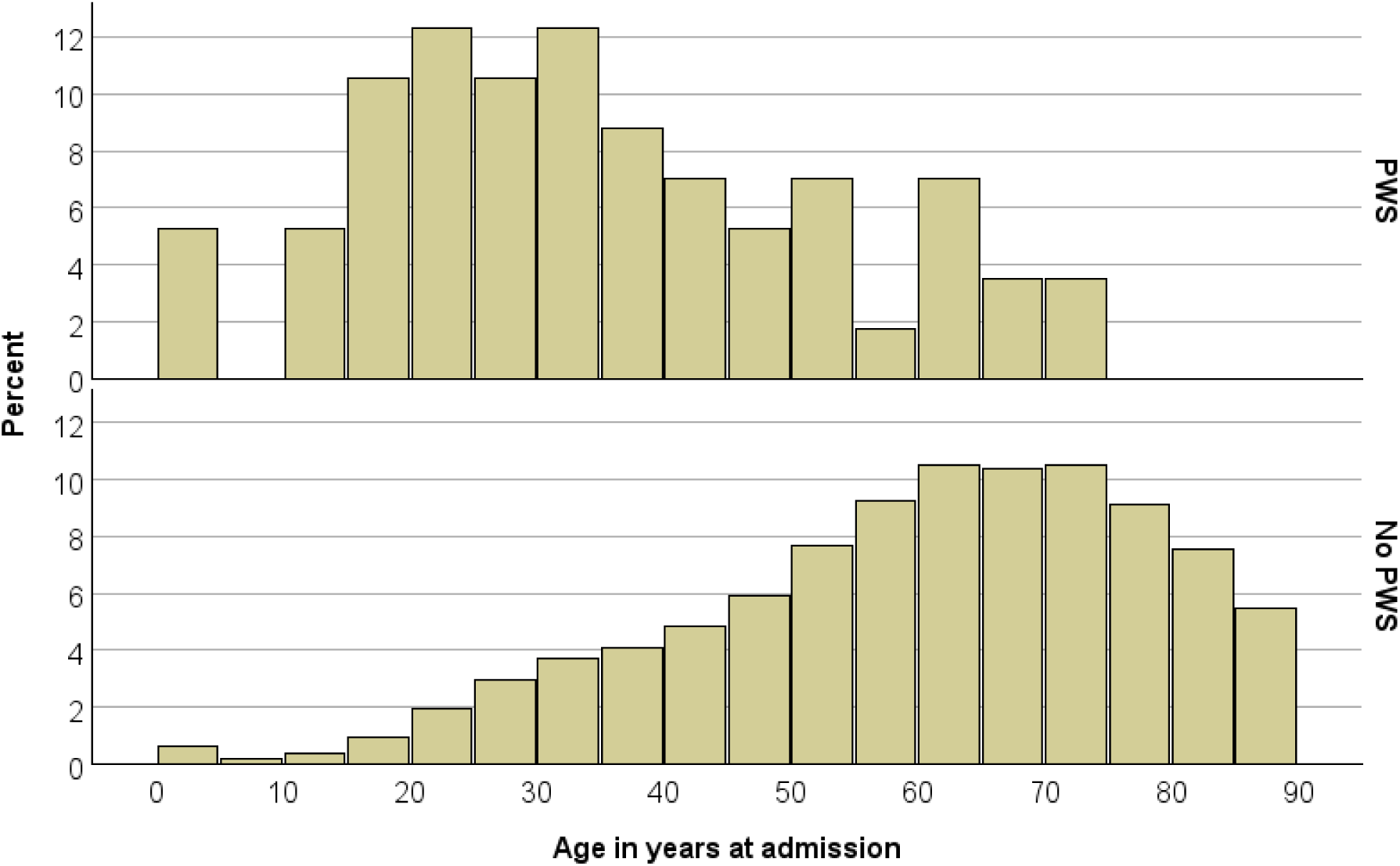
age distribution of hospitalizations for patients with COVID-19 infection with a diagnosis of PWS (top) and without a PWS diagnosis (bottom).

**Table 1:**
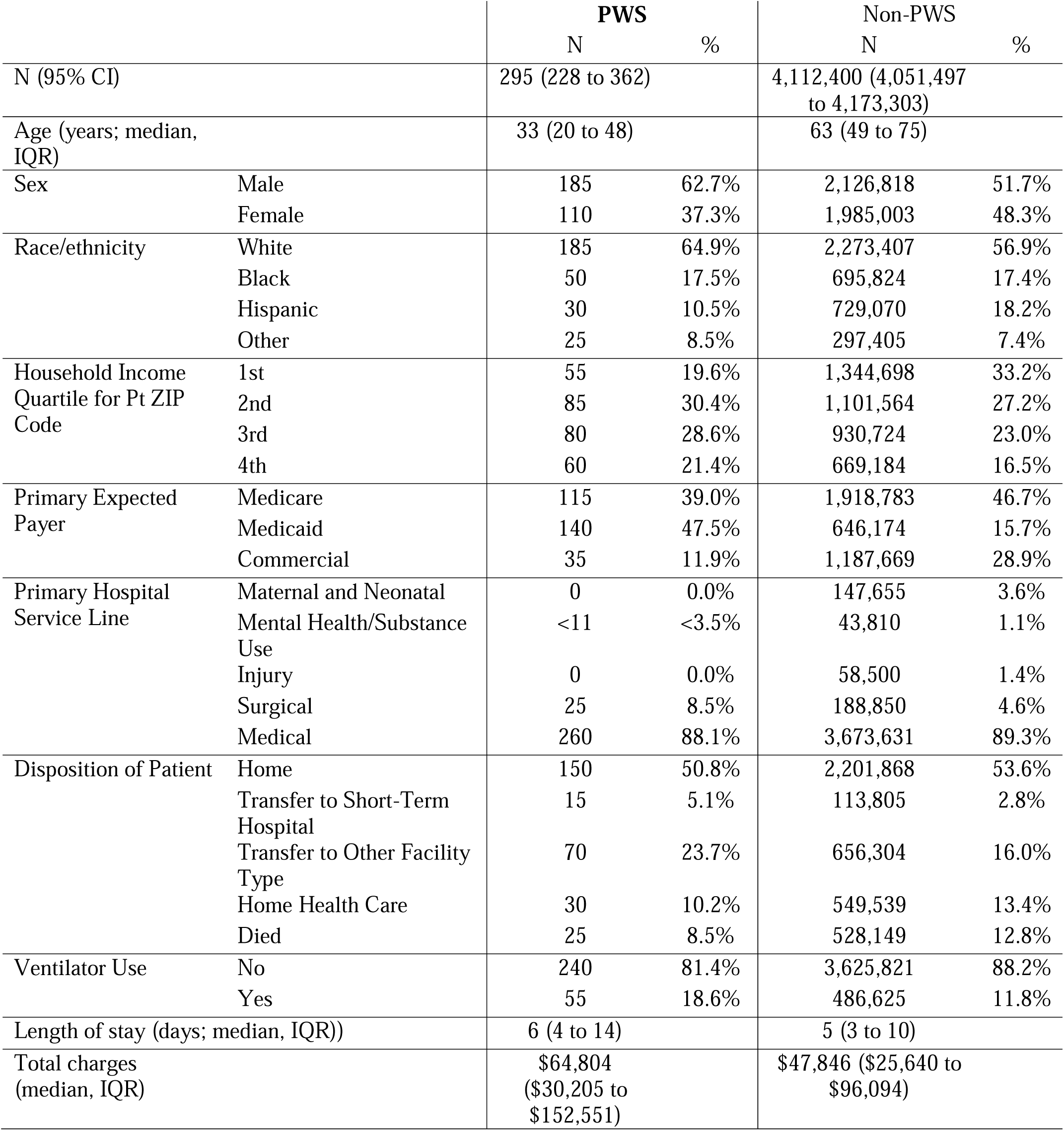
demographics of patients hospitalized with COVID-19 infections with and without a diagnosis of PWS.

Among baseline comorbidities, obesity codes were present in 47.5% of PWS hospitalizations compared to 28.4% of non-PWS hospitalizations, while chronic pulmonary disease was present in 23.7% of PWS hospitalizations and 21.4% of non-PWS hospitalizations. Baseline comorbidities are listed in Table 2.

**Table 2:** baseline comorbidities of patients hospitalized with COVID-19 infection with and without a diagnosis of PWS.

| Comorbidity | *PWS* |  | *No PWS* |  |
| --- | --- | --- | --- | --- |
|  | N | % | N | % |
| Acquired immune deficiency syndrome | 0 | 0.0% | 20,085 | 0.5% |
| Alcohol abuse | 0 | 0.0% | 102,230 | 2.5% |
| Autoimmune conditions | 20 | 6.8% | 133,730 | 3.3% |
| Cancer | 0 | 0.0% | 182,215 | 4.4% |
| Dementia | 0 | 0.0% | 365,725 | 8.9% |
| Depression | 25 | 8.5% | 436,400 | 10.6% |
| Diabetes without chronic complications | 45 | 15.3% | 493,539 | 12.0% |
| Diabetes with chronic complications | 90 | 30.5% | 1,012,204 | 24.6% |
| Drug abuse | 0 | 0.0% | 100,275 | 2.4% |
| Hypertension, complicated | 35 | 11.9% | 1,004,604 | 24.4% |
| Hypertension, uncomplicated | 105 | 35.6% | 1,483,158 | 36.1% |
| Chronic pulmonary disease | 70 | 23.7% | 879,984 | 21.4% |
| Obesity | 140 | 47.5% | 1,166,144 | 28.4% |
| Peripheral vascular disease | 5 | 1.7% | 164,025 | 4.0% |
| Hypothyroidism | 65 | 22.0% | 514,039 | 12.5% |
| Other thyroid disorders | 0 | 0.0% | 48,260 | 1.2% |

Among COVID hospitalizations for individuals with PWS, 18.6% involved the use of mechanical ventilation, compared to 11.6% of hospitalizations for individuals without PWS. In-hospital death occurred in 8.5% of hospitalizations for individuals with PWS compared to 12.8% of hospitalizations for those without PWS. Among patients who died, median age was 30 years for PWS patients and 72 years for those without PWS. Overall length of stay was a median of 6 days (IQR: 4 to 14) for patients with PWS compared to 5 days (IQR: 3 to 10) for those without PWS. Median hospital charges were $64,804 (IQR: $30,205 to $152,551) for individuals with PWS compared to $47,846 (IQR: $25,640 to $96,094) for individuals without PWS.

To estimate the effect of PWS diagnosis on hospital outcomes, AIPW was performed with PWS diagnosis (yes/no) as a binary exposure variable with age, sex, race, primary hospital service line, and year as covariates (Table 3). In these models, PWS diagnosis was associated with a 7.43 day increase in hospital length of stay and a $80,126 increase in hospital charges.

**Table 3:**
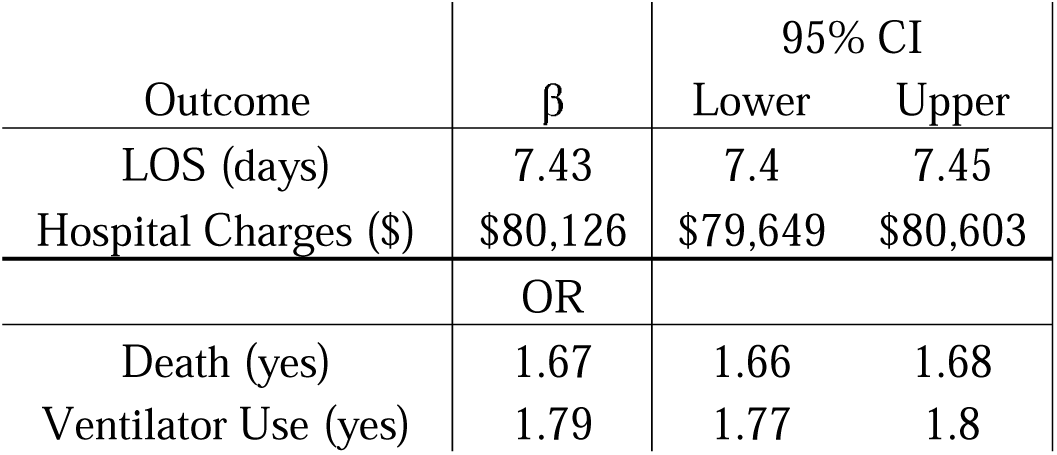
AIPW models of the causal effect of a PWS diagnosis (independent variable) on outcomes of hospitalizations involving COVID-19 infection, with age, sex, race, primary hospital service line, and year as covariates.

| Outcome | $\beta$ | 95% CI | |
| --- | --- | --- | --- |
|  |  | Lower | Upper |
| LOS (days) | 7.43 | 7.4 | 7.45 |
| Hospital Charges (\$) | \$80,126 | \$79,649 | \$80,603 |
|  | OR |  |  |
| Death (yes) | 1.67 | 1.66 | 1.68 |
| Ventilator Use (yes) | 1.79 | 1.77 | 1.8 |

PWS was associated with a higher odds of mechanical ventilation utilization, with an odds ratio (OR) of 1.79. PWS diagnosis was additionally associated with a higher odds of death during hospitalization (OR 1.67).

## Discussion

In 2020 and 2021, an estimated 295 (95% CI: 228 to 362) hospitalizations for patients with PWS and COVID-19 occurred in the United States. In adjusted models, compared to hospitalizations for patients without PWS, those with PWS had substantially longer hospital length of stay and hospital charges as well as increased odds of requiring mechanical ventilation or dying during hospitalization. Together, these results demonstrate the substantial clinical impact of COVID-19 infection on hospitalization outcomes for individuals with PWS as well as the increased cost and duration of these hospitalizations.

Patients with PWS hospitalized with COVID-19 had a median age of 33 years, compared to 63 years for those without PWS. As age is substantially associated with COVID-19 mortality [Elo et al., 2022], older median age likely explains much of the higher crude mortality observed in non-PWS hospitalizations. Among patients who died, median age for patients with PWS was 42 years younger than for patients without PWS, highlighting the early in-hospital mortality for individuals with PWS hospitalized with COVID-19 compared to those without PWS. This suggests that, contrary to prior survey studies, the clinical course of COVID-19 is not milder for patients with PWS in the subset of individuals who are ill enough to require hospitalization.

These inpatient data cannot, however, inform the overall clinical course of COVID-19 infection in individuals with PWS as it is possible that the overall severity of infectious symptoms in PWS is equal, less severe, or more severe than in the general population. A hypothesized mechanism of improved innate immunity in individuals with PWS resulting in lower overall viral disease severity remains a possibility [Viardot et al., 2010]. These data do suggest, however, that severe COVID-19 infection resulting in hospitalization and early death remains possible in the context of PWS as may be expected from the baseline impaired respiratory function in such individuals.

As a result, individuals with PWS should be considered at high risk for severe COVID-19, and patients and families should continue to utilize strategies, including vaccination, to reduce the risk of severe complications of infection.

The overall number of COVID-19-associated in-hospital deaths in 2020 and 2021 captured by the NIS was 528,149, which is comparable to estimates of in-hospital deaths during the same time period (579,199) from the National Center for Health Statistics of the US Centers for Disease Control and Prevention (CDC) using different data reporting methodology [COVID-19 Provisional Counts - Weekly Updates by Select Demographic and Geographic Characteristics, 2023]. The CDC data counted a total of 848,943 COVID-19-associated deaths in all settings during this period, meaning that 32% of COVID-19 deaths occurred outside of the inpatient setting. It is unclear if this ratio for the general population applies to individuals with PWS, or if a lower or higher fraction of deaths of individuals with PWS occurred inpatient and is captured by this analysis.

Baseline comorbidities including obesity, pulmonary disease, and vascular disease were common in both PWS and non-PWS hospitalizations. While coding for obesity is known to be poor in the NIS overall [Al Kazzi et al., 2015], the rate of obesity in PWS patients in this sample (47.5%) is higher than that described in survey studies assessing for COVID-19 among PWS patients [Coupaye et al., 2021; Whittington et al., 2022]. As a result, the effects of COVID-19 on hospitalization outcomes may be mediated by these clinical variables. Due to small sample size we were unable to perform formal mediation analysis of covariates, but with larger samples this could inform the mechanism of increased disease severity among individuals with PWS and inform strategies for mitigating a portion of this risk.

The COVID-19 pandemic had complex effects on individuals with PWS resulting from its significant impact on educational, social, and healthcare access. Evidence suggests that individuals with PWS can benefit from strict dietary control in specialist group home settings [Mastey Ben-Yehuda et al., 2024], with pandemic-related closures affecting access to these placements in some regions. Survey studies in Germany [Mohr et al., 2021], France [Mosbah et al., 2021], and Italy [Baietto et al., 2023] have indicated conflicting impacts of lockdowns on clinically-relevant outcomes including weight and mental health, which may be explained by differences in lockdown protocols and baseline treatment in different regions. As a result, the optimal care for individuals with PWS for any future pandemics must balance potential risks from disruptions of routine with potential risks of infection to PWS patients who may be at high risk of severe complications.

By utilizing a large, nationally-representative data source covering hospitalizations across the US regardless of payor, this study provides evidence of clinical care for individuals with PWS without requiring active participation of study participants. This enhances the generalizability of these findings. Prior validation studies have demonstrated excellent reliability of the administrative claim code for COVID-19[Zhong et al., 2021], which means that this study should capture most episodes of inpatient care for COVID-19 infection during the study period, with 2021 data being the most recent that are available.

One key limitation of the study is potential errors in coding for PWS using the Q87.11 code, which came into clinical use in 2019. As a result, any errors in PWS diagnosis or in coding for the disorder would bias results here. As a result, validation of the Q87.11 code is a critical next step in administrative claims research for PWS. Additionally, the single Q87.11 code does not distinguish between genetic subtypes of PWS and so this study is unable to inform potential clinical differences between subgroups of PWS. Furthermore, the NIS surveys hospitalizations and not individual patients, and so if an individual is treated in more than one hospital during the study period or is re-hospitalized, that individual may appear more than once in the NIS data.

Moreover, these results are from 2020 and 2021, and since that time the circulating COVID-19 virus has mutated and there have been changes in population-level immunity caused by vaccination and prior infection. As a result, further and ongoing research is needed to provide optimal treatment recommendations for individuals with PWS as the COVID-19 pandemic continues to evolve. Finally, although AIPW is designed as a means of estimating average causal effects from observational data, we note the large literature on the imperfection of replication of casual effects found in randomized trials through quasi experimental methods applied to observational data [Hemkens et al., 2016; Benson and Hartz, 2000; Moss et al., 2019; Wang et al., 2023].

## Conclusion

In 2020 and 2021 there were 295 (95% CI: 228 to 362) hospitalizations for patients with PWS and COVID-19 in the United States. Compared to hospitalizations for those without PWS, individuals with PWS had longer length of stay, higher hospital charges, higher odds of ventilator use, and higher mortality. These results highlight severe manifestations of COVID-19 infection in individuals with PWS, warranting continued protective measures and vaccination efforts. Further research is needed to validate coding for PWS and assess the impact of evolving COVID-19 variants and population immunity on this vulnerable population.

## Data Availability

Publicly available datasets were analyzed in this study. The dataset analyzed for this study can be obtained from the Healthcare Cost and Utilization Project (HCUP), Agency for Healthcare Research and Quality at:
https://www.hcup-us.ahrq.gov/db/nation/nis/nisdbdocumentation.jsp

